# Matching Patients to Clinical Trials using LLaMA 2 Embeddings and Siamese Neural Network

**DOI:** 10.1101/2024.06.28.24309677

**Authors:** Shaika Chowdhury, Sivaraman Rajaganapathy, Yue Yu, Cui Tao, Maria Vassilaki, Nansu Zong

**Affiliations:** Mayo Clinic, Department of AI and Informatics Research, Rochester, MN, US; Mayo Clinic, Department of Quantitative Health Sciences, Rochester, MN, US

## Abstract

Patient recruitment is a key desideratum for the success of a clinical trial that entails identifying eligible patients that match the selection criteria for the trial. However, the complexity of criteria information and heterogeneity of patient data render manual analysis a burdensome and time-consuming task. In an attempt to automate patient recruitment, this work proposes a Siamese Neural Network-based model, namely Siamese-PTM. Siamese-PTM employs the pretrained LLaMA 2 model to derive contextual representations of the EHR and criteria inputs and jointly encodes them using two weight-sharing identical subnetworks. We evaluate Siamese-PTM on structured and unstructured EHR to analyze their predictive informativeness as standalone and collective feature sets. We explore a variety of deep models for Siamese-PTM’s encoders and compare their performance against the Single-encoder counterparts. We develop a baseline rule-based classifier, compared to which Siamese-PTM improved performance by 40%. Furthermore, visualization of Siamese-PTM’s learned embedding space reinforces its predictive robustness.

## Introduction

Randomized clinical trials (RCT) play a vital role in clinical research as they are conducted to assess the efficacy of new treatments or interventions. Marked by their high failure rates, however, clinical trials remain the primary bottleneck in the drug development pipeline, with more than two-thirds of Phase II compounds failing to advance to Phase III^1^. One of the main drivers of an unsuccessful clinical trial pertains to the problem of patient recruitment whereby enough eligible participants cannot be accrued for the study on time. It is estimated that 86% of clinical trials fail to meet the patient recruitment goals^2^, resulting in 19% of trials being terminated^3^. The foremost challenge underlying the recruitment process is the difficulty in matching study-specific inclusion and exclusion criteria to patient records. Given the magnitude and diversity of the electronic health records (EHRs) data with a mix of structured and unstructured patient information, manually reviewing and analyzing them based on predefined selection criteria can be a laborious task. In addition, for more extensive and complex eligibility criteria, filtering patient records based on simple keyword matches is unproductive. Therefore, there exists a critical need to develop computational models that can automatically process EHR and criteria data to accurately gauge patients’ eligibility for cohort selection, toward achieving optimized clinical trial design.

Previous works in patient-trial matching encompass rule-based^4-6^, machine/deep learning-based^7,8^, and hybrid methods^9,10^. Rule-based methods rely on carefully crafted heuristics to describe the selection criteria. Machine/deep learning methods, by contrast, are capable of automatically learning criteria-related patterns from the data. While hybrid methods employ rule-based method as an auxiliary step to either perform manual feature extraction or post-processing for a machine learning-based method. The major limitations of rule-based methods are the taxing rule engineering process and the lack of generalizability to new datasets. Traditional machine learning methods are capable of automatic data modeling, albeit human expertise is usually required for selecting the trial-specific feature set. Deep learning methods, on the other hand, are data-driven and can learn the optimal feature set automatically from the raw data, allowing it to select the most eligible patients for any clinical trial with minimal human intervention.

The application of deep learning for patient-trial matching is faced with the challenge of the limited data availability. Clinical trials test the safety and effectiveness of new treatment, so it is plausible that a limited patient population is generally exposed to it. Taking into account the data-hungry nature of deep learning techniques and given the resource-scarce setting of the patient-trial matching task, this work proposes a **<u>p</u>**atient-**t**rial **<u>m</u>**atching framework based on the **<u>Siamese</u>** Neural Network architecture, namely **Siamese-PTM**. Siamese-PTM employs two subnetworks with the same configuration and weights to encode the patient EHR vector and the selection criteria vector in parallel. The pairwise feature learning generates distributed representations of the EHR and criteria data that more precisely capture the semantic interactions between them to facilitate accurate predictions of patient eligibility. As a result of the paired inputs ingested by Siamese-PTM, the number of training samples is implicitly increased, allowing it to overcome the deficiency of training data and consequently generalize well on new patient-criteria inputs. We further enrich Siamese-PTM’s encoding capability by utilizing the pretrained LLaMA 2^11^ model to initialize the embeddings of the EHR and criteria inputs.

We evaluate Siamese-PTM’s performance on structured and unstructured EHR data to investigate their individual and collective predictive value in the patient-trial matching task. We analyze the effect of fine-grained (token-level) and coarse-grained (document-level) pretrained LLaMA 2 embeddings on Siamese-PTM’s generalization. We perform comparative quantitative analysis that explored a variety of neural network architectures for Siamese-PTM’s encoders and compared their performance against the Single-encoder counterparts. We develop a baseline rule-based classifier, compared to which Siamese-PTM significantly improved performance by 40% (p-value=0.006). To further demonstrate the representation strength of the embedding vectors learned by Siamese-PTM, a qualitative evaluation is carried out in the form of cluster analysis^12^.

## Methods

### Study design and data

We use five clinical trials (NCT02008357, NCT04468659, NCT02669433, NCT01767909, NCT02565511) from ClinicalTrials.gov focused on cognitive disorders, such as Alzheimer’s disease and Dementia with Lewey Bodies. For each trial, the selection criteria data consist of inclusion and exclusion statements extracted from the “Eligibility Criteria” section of that trial. Cohort definition per trial was formed by applying the structured query language (SQL) to the EHR data according to the respective selection criteria, resulting in a total of 180 patients. Out of the 180 patients, 50 were eligible (“success”) and the remaining 130 did not satisfy the trial’s criteria (“fail”). The EHR data was collected from Mayo Clinic’s United Data Platform (UDP) and included structured, unstructured and demographic patient data. The structured EHR covers six types of clinical events (diagnosis, medication, allergy, family history of medical condition, lab tests, and admission (e.g., reason for visit)); the unstructured EHR comprises radiology reports; and demographics includes patient’s age and gender. We consider five criteria-EHR pairwise input combinations for evaluation: (1) *criteria + unstructured EHR*, (2) *criteria + structured EHR*, (3) *criteria + unstructured and demographics EHR*, (4) *criteria + structured and demographics EHR* and (5) *criteria + unstructured, structured and demographics EHR*.

### Problem formulation

The patient-trial matching task in this work is considered as a binary classification problem, which can be formulated as follows: Given <EHR, Criteria> pairs *I* = {<*E*_*1*_, *C*>,….,<*E*_*i*_, *C*>,….,<*E*_*P*_, *C*>} across *P* patients, where *E*_*i*_ = {*e*_*i1*_, *e*_*i2*_,….,*e*_*id*_} is the EHR data of the *i*-th patient composed of *d* tokens and *C* = {*c*_*1*_,….,*c*_*j*_,….,*c*_*m*_} is the sequence of *m* criteria statements in our study, with criterion *c*_*j*_ containing *n* tokens {*c*_*j1*_,….,*c*_*jn*_}, the goal is to predict whether the *i*-th patient meets the criteria (“success”) or not (“fail”).

### Overview of Siamese-PTM

Provided that each training instance in the patient-trial matching task is a <EHR, Criteria> pair, the canonical approach to integrating the two types of information is by performing early fusion^**13**^. As depicted in Figure 1 (a), the early fusion approach performs the integration on the input side by combining the <EHR, Criteria> vectors to a unified input vector and trains a single encoder to perform the prediction. Note that the merging of the pairwise vectors can take place using any fusion operation (e.g., concatenation). However, owing to the fact that the semantic similarity between the EHR and criteria could exist beyond the surface level, early fusion fails to capture the interactions between the two data as a single encoder operates on the integrated data. To address this shortcoming, this work adopts the intermediate fusion^13^ approach that performs the integration of the data during the training phase by adopting the Siamese Neural Network architecture, namely Siamese-PTM. As depicted in Figure 1 (b), Siamese-PTM first employs two identical encoders to learn feature representations of the EHR and criteria data separately, which are subsequently integrated and passed through the classification layer. By sharing weights between the two encoder modules and training them jointly, Siamese-PTM is able to learn correlations between the two types of information. In doing so, the semantic relevance between the <EHR, Criteria> pair can be accurately modeled and patients matching the eligibility criteria can be effectively identified.

**Figure 1.**
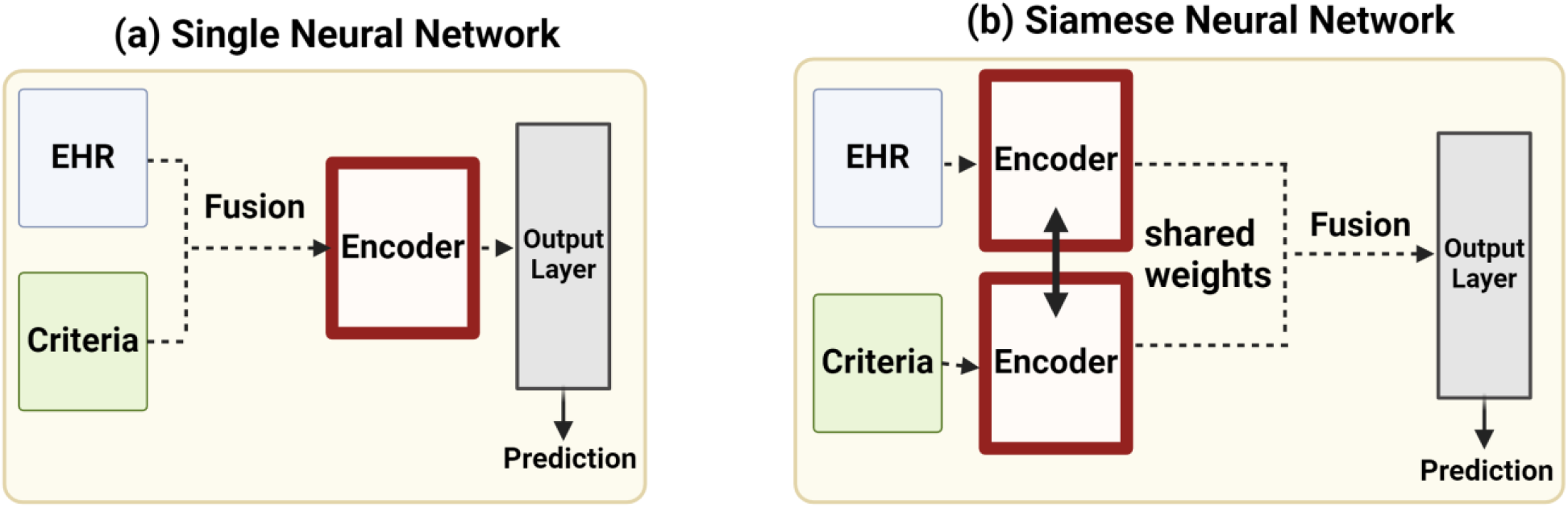
Overview of the Single Neural Network model with one encoder and Siamese Neural Network with two encoders.

The encoded representations of the <EHR, Criteria> pair learned by the dual encoders in Siamese-PTM are then fed into an output layer, which is implemented as a linear layer with one hidden unit followed by sigmoid activation. In order to fuse the two encoded representations into a single vector to perform classification, we experiment with the following techniques: concatenation and element-wise addition, multiplication and mean.

### LLaMA 2 pretrained embeddings

Large language models constitute Transformer-based models delineated by their considerable scale and pretraining on large corpora of unlabeled data. LLaMA 2 is an emerging autoregressive model that has substantial world knowledge ingrained in its 7 billion trainable parameters by learning language representation in a self-supervised way. This capability allows it to capture complex contextual relationships and improve the performance of various downstream tasks. We utilize the pretrained LLaMA 2^14^ model to derive the contextualized representations of the <EHR, Criteria> pair as input features for Siamese-PTM. To capture the semantic nuances at different granularity, we evaluate Siamese-PTM on fine-grained and coarse-grained LLaMA 2 embeddings. In the fine-grained analysis, each token in the EHR *E*_*i*_ and criteria *C* sequences is represented with its token-level embedding extracted from LLaMA 2. As a result, fine-grained analysis enables Siamese-PTM to model the sequential relation between the token representations within <EHR, Criteria>. In Coarse-grained analysis, on the other hand, the aggregated contextual representations outputted by LLaMA 2 for the EHR and criteria are inputted directly into Siamese-PTM. Note that in this case, we consider the whole EHR sequence *E*_*i*_ as a single text corresponding to the document, while each criterion statement *c*_*j*_ in *C* is considered as independent text and mapped to its LLaMA 2 embedding, which is then mean-pooled to represent *C* at the document-level. So, coarse-grained analysis summarizes the overall semantics of the EHR and criteria data into single contextual representations, which are then used to initialize Siamese-PTM’s learning.

### Encoder

The siamese encoder architecture used in coarse-grained analysis is a multilayer layer perceptron (MLP)^15^ and for fine-grained analysis we experiment with a wide range of neural networks, which include the long short-term memory network (LSTM)^16^, gated recurrent unit (GRU)^17^ network, their bidirectional variants (i.e., Bi-LSTM^18^, Bi-GRU^19^), their attention variants^20,21^ (i.e., LSTM-ATT, GRU-ATT, Bi-LSTM-ATT, Bi-GRU-ATT), convolutional neural network (CNN)^22^ and CNN-LSTM^23^.

#### MLP

Also referred to as a feed forward neural network, MLP consists of one or more hidden layers of neurons (hidden units) with non-linear activation (ReLU^24^) and full connectivity between them. We apply batch normalization^25^ and dropout^26^ layers as regularization techniques after each hidden layer. We vary the number of hidden layers, number of hidden units, and dropout rate and analyze their effect on Siamese-PTM’s performance.

#### LSTM

It contains special memory cells and uses a gating mechanism featuring three gates (input, forget, output) to control the information flow within the cells, allowing LSTM to tackle the vanishing gradient problem inherent in recurrent neural networks (RNN)^27^ and capture long-term dependencies. We use LSTM with one hidden layer of 50 hidden units and a dropout layer with dropout rate of 0.5 based on preliminary hyperparameter tuning.

#### GRU

Similar to LSTM, but GRU has a simplified architecture with two gates (update and reset) to control the information flow and as a result fewer parameters. We use the same hyperparameter settings as LSTM.

#### Bidirectional variants

The Bi-LSTM and Bi-GRU models are composed of the forward LSTM/GRU and backward LSTM/GRU to capture dependencies from both directions. We use the same hyperparameter settings as the vanilla variants.

#### Attention variants

We apply self-attention mechanism to the hidden states of the LSTM/GRU variants and compute the output as the attention-weighted sum of the hidden states.

#### CNN

We apply 1-d convolving filter across the embedding dimension to extract feature vectors and perform mean-pooling to aggregate them into a single vector.

#### CNN-LSTM

This is a hybrid model that first extracts feature vectors leveraging a CNN and passes them through an LSTM for sequential modeling.

### Loss function

To train Siamese-PTM for the patient-trial matching task, we experimented with three loss objectives as below. For BCE and WBCE, Siamese-PTM’s learned representations are fused and passed through the output layer to get the predicted probability. While for CL, we compute the cosine similarity between the learned representations.

#### Binary cross-entropy (BCE)

Quantifies Siamese-PTM’s performance with predicted probability ranging from 0 to 1. The loss tends to increase if the output probability *ŷ* is different from the ground truth label *y* (i.e., fail or success). Mathematically, it is defined as below.

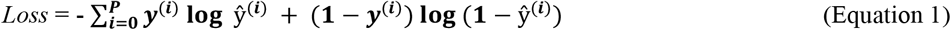

#### Weighted binary cross-entropy (WBCE)

It is based on BCE but addresses the class imbalance in the dataset by introducing the weight β to penalize misclassifications of the minority class more.

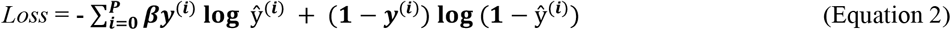

#### Contrastive loss (CL)

It penalizes the encoder based on the eligibility label *y* of the *i*-th <EHR, Criteria> pair such that similar feature embeddings are learned for the EHR and criteria inputs with the “success” label and dissimilar feature embeddings are learned for pairs with the “fail” label. Here, *d*_*w*_ is the cosine similarity between the learned feature embeddings of the EHR and criteria inputs and *margin* is a hyperparameter which is maximized for positive samples.

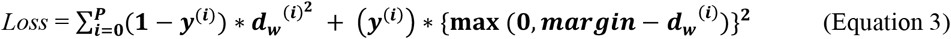

## Results

### Model evaluation

Models were trained, validated and tested in a five-fold cross-validation. We report performance in terms of the mean F1 score averaged across the five-fold results. We used Adam optimizer to train each model for 50 epochs with a learning rate of 0.005.

### Baseline rule-based results

For comparison, we develop a baseline rule-based patient-trial matching classifier where criteria-based rules crafted in the form of SQL query are utilized to retrieve specific clinical features from EHR, guided by expert clinician knowledge. The retrieved features are subsequently fed as input into a standard machine learning model to perform binary classification. We experiment with three different models for evaluation as shown in Figure 2 (a). The results suggest that the best performing baseline classifier is Random Forest, followed by SVM, while Logistic Regression performs the worst. On inspecting the per class generalization, however, SVM outperforms Random Forest in accurately predicting the eligible patients that match the criteria (F1-Pos).

**Figure 2.**
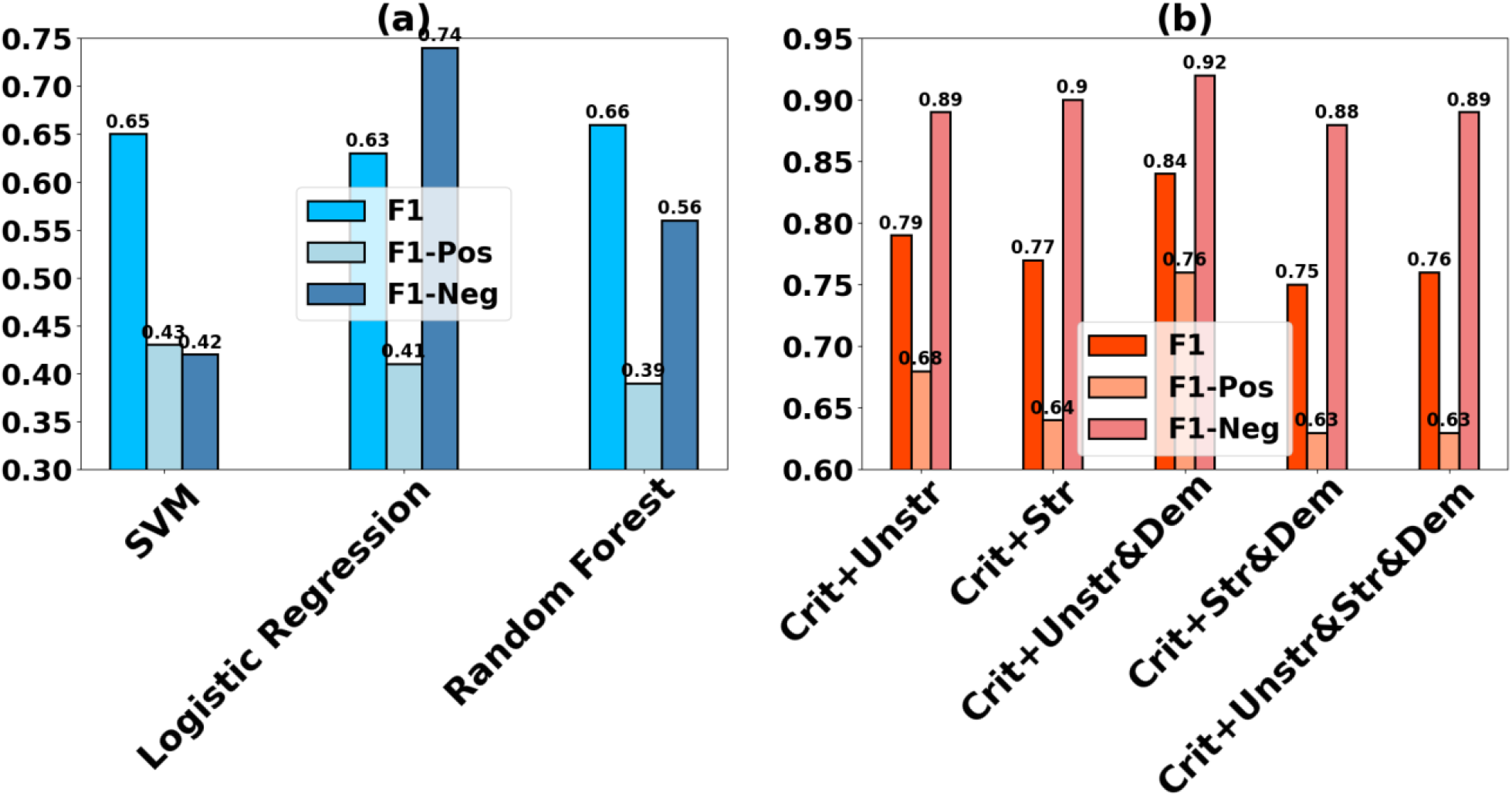
(a) Baseline rule-based performance evaluated with traditional machine learning models. (b) Effect of different input combinations on Siamese-PTM’s performance.

### Importance of EHR data type

Figure 2 (b) examines the individual and collective influence of the structured and unstructured EHR data on Siamese-PTM’s performance in the coarse-grained setting. We observe that the most informative data source is the unstructured narrative data (i.e., radiology reports), both as a standalone feature set (criteria + unstr) as well as coupled with the demographics feature (criteria + unstr & dem), leading to F1 scores of 0.79 and 0.84 respectively. The counterpart structured input combinations (criteria + str, criteria + str & dem), however, underperform with performance gaps of 3% and 11%, respectively, in comparison to the corresponding unstructured input combinations. As a result, the performance on integrating both unstructured and structured EHR into the input (criteria + unstr & str & dem) is balanced by the informativeness of each data type (F1 of 0.76).

### Coarse-grained analysis

To gain a deeper understanding of Siamese-PTM’s predictive capabilities in the coarse-grained analysis, we change the MLP encoder’s hyperparameter settings with respect to the projection dimension, number of hidden layers, dropout rate and fusion approach and analyze the effect of each on the model’s performance. The projection dimension corresponds to the output dimension of the encoded representation learned by Siamese-PTM and is varied as {5, 10, 30, 50, 64, 100}. The results shown in Figure 3 (a) suggest that projecting to a higher dimension generally leads to a better performance. Specifically, the best performance was achieved with dimension 64 and worst performance with dimension 5. However, increasing the dimension further (100) degrades performance, possibly due to overfitting caused by the small dataset vs. large feature dimensionality ratio (the curse of dimensionality). Then, to inspect the learning ability of an increasingly deeper model, we stack the MLP with 1, 2, 3 or 4 hidden layers. The declining performance in Figure 3 (b) alludes that Siamese-PTM overfits more as we increase the model complexity with an additional hidden layer every time. The overfitting could potentially be caused by trying to fit a complex model on a rather small sized dataset. Dropout is performed to prevent overfitting by randomly dropping neurons from the hidden layer, as specified by the dropout rate. We experiment with the following dropout rates {0.1, 03, 0.5, 0.8, 0.9} as shown in Figure 3 (c), among which 10% dropout is optimum. Generally, a high dropout rate is known to hurt the performance of models trained on small datasets^26^ and our results align with this finding to some extent, although it is not conclusive. Lastly, Figure 3 (d) presents Siamese-PTM’s performance with different fusion approach that merge the learned representations outputted by the dual encoders into a single vector. Among them, concatenation performs the best, followed by addition obtaining competitive result, while mean and multiplication perform relatively worse.

**Figure 3.**
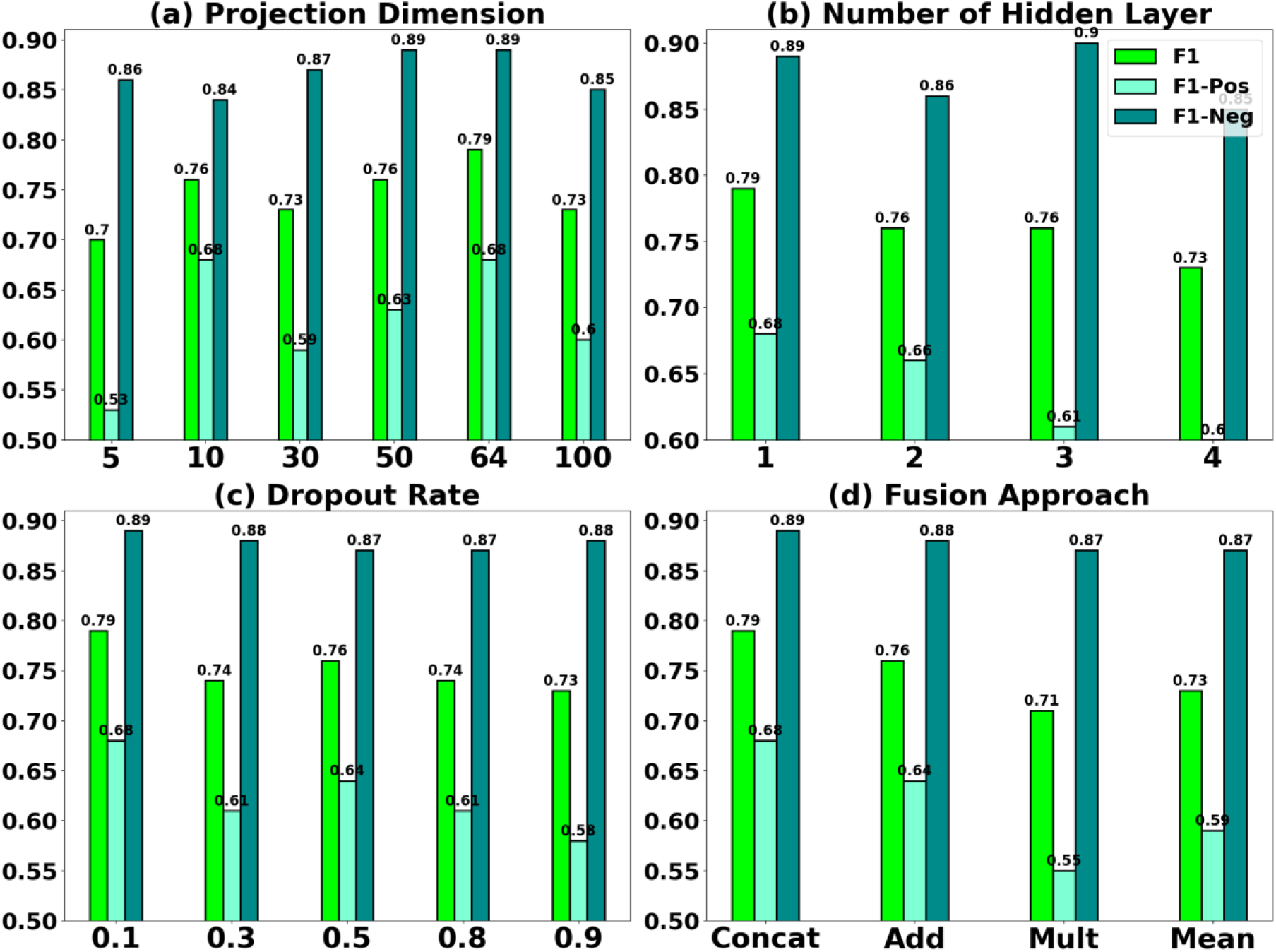
Coarse-grained analysis of Siamese-PTM evaluated on (a) projection dimension, (b) number of hidden layers, (c) dropout rate and (d) fusion approach.

### Fine-grained analysis

Figure 4 (a) reports the fine-grained performance of Siamese-PTM on the unstructured EHR data having the encoder replaced with different neural network models. Among the encoders, LSTM ranks best with an F1 score of 0.92. This deep learning performance is much above the rule-based Random Forest model’s performance (0.66 F1), in particular by a statistically significant 40% performance gain (p-value = 0.006). Other LSTM variants, namely GRU and Bi-LSTM, perform comparably to each other (0.86 F1), while Bi-GRU performs relatively worse (0.81 F1). In general, the attention-augmented variants improve performance, with the Bi-LSTM-ATT (0.88 F1) and GRU-ATT (0.91 F1) models achieving 2% and 6% respective boosts in performance compared to their vanilla variants. While CNN (0.88 F1) performs comparably to the LSTM-based attention variants and CNN-LSTM’s result (0.86 F1) is comparable to that of Bi-LSTM and GRU.

**Figure 4.**
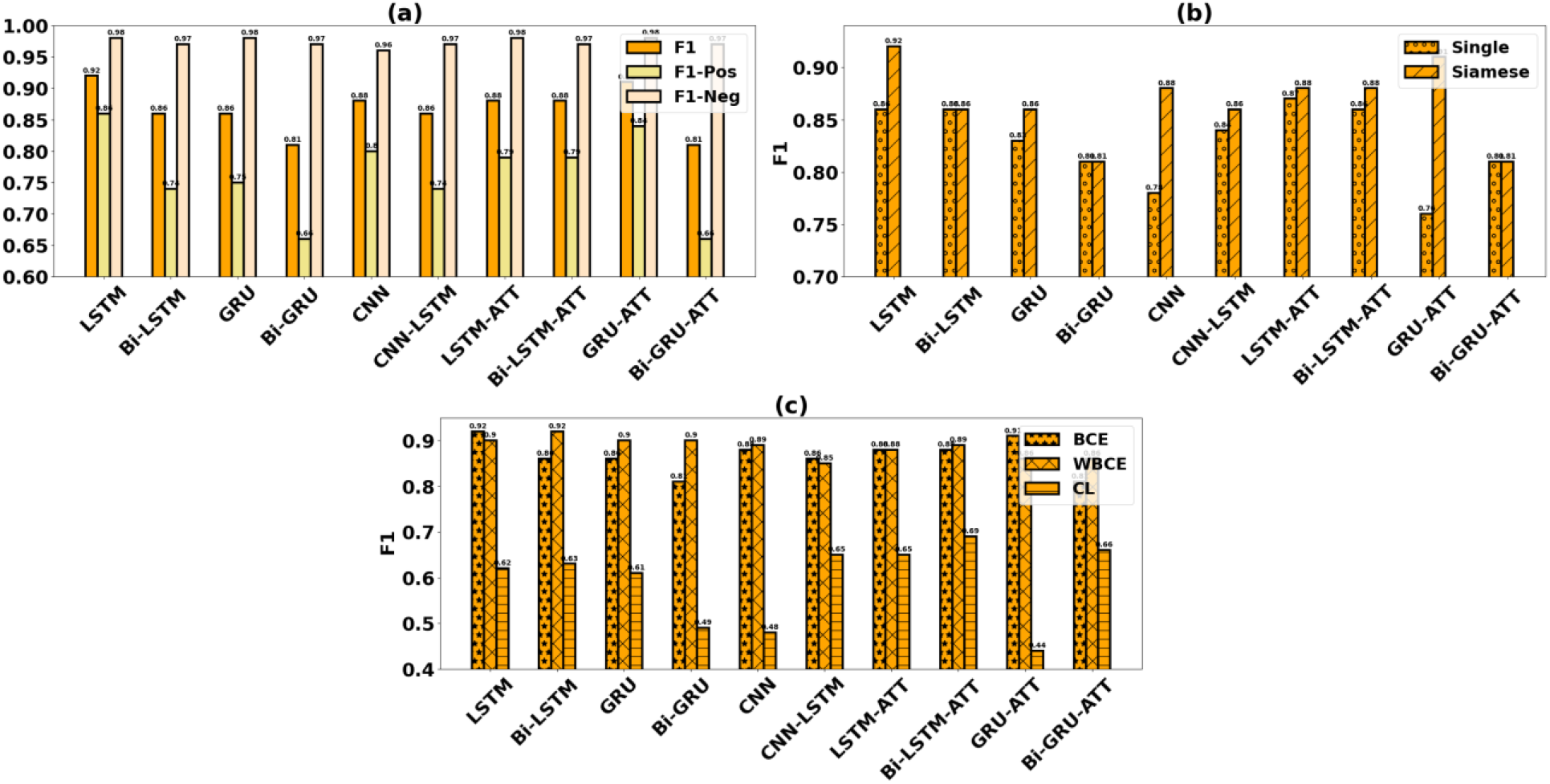
Fine-grained analysis of Siamese-PTM (a) evaluated with different encoders, (b) compared against Single-encoder models and (c) evaluated with different loss functions.

To justify the adoption of a Siamese Neural Network-based architecture for the patient-trial matching task, in Figure 4 (b) we compare the proposed Siamese-PTM’s performance with the corresponding Single-encoder models. The results strongly assert Siamese-PTM’s superiority as it is able to outperform seven out of the ten counterpart Single-encoder models, while performing comparably with the remaining three.

In Figure 4 (c), we analyze Siamese-PTM’s performance evaluated with different loss objectives. As there exists an imbalance in our class distribution, employing the weighted variant of the binary cross-entropy loss (i.e., WBCE) helps to improve generalization of six out of the ten encoders compared to the standard BCE loss. This is attributed to WBCE assigning higher penalty to the misclassifications of the minority class (“success”), while improving Siamese-PTM’s sensitivity and thus its overall performance. Contrastive loss (CL), on the other hand, relatively underperforms for this task. Hypothesizing, in this study we reduce a patient-trial matching task to a binary classification problem; however, CL being a metric-based loss, is designed to model the similarity between inputs by learning similar feature embeddings for inputs belonging to the same class and dissimilar embeddings for inputs belonging to different class. As a result of this difference in task objectives and because the patient-trial inputs do not directly map to similar semantics, CL is not able to perform at par with the alternative loss functions BCE and WBCE.

### Cluster analysis

We perform cluster analysis of the patient-trial representations learned by Siamese-PTM (i.e., fused representations right before passing to the output layer) after dimensionality reduction with principal component analysis (PCA)^28^ and t-distributed stochastic neighbor embedding (tSNE)^12^ techniques. We visualize the results derived from different training epochs in Figure 5. The visualization shows that similar patients are grouped together matching the label of their trial eligibility (“success” or “fail”). The distinct clusters demonstrate that Siamese-PTM is able to capture semantics that align with the domain knowledge and further solidifies its robust representation learning capability for the patient-trial matching task.

**Figure 5.**
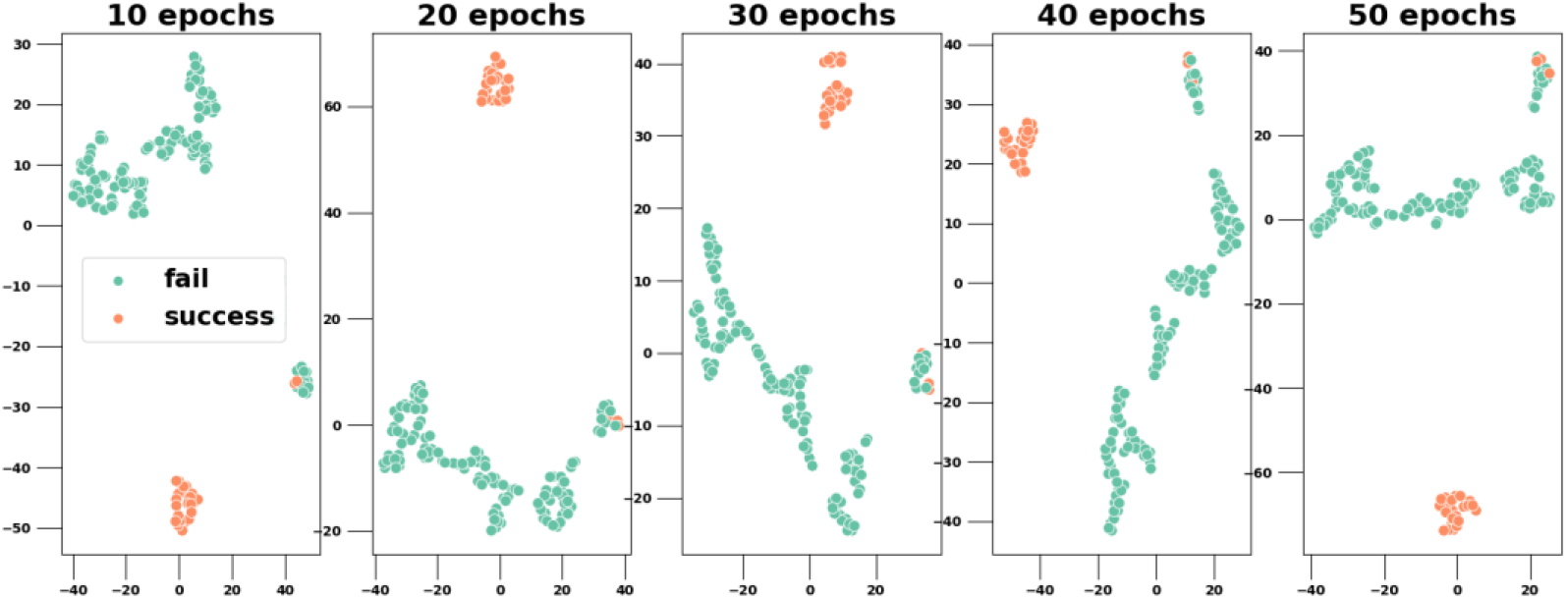
Cluster analysis of Siamese-PTM’s learned representations across different epochs. Each dot corresponds to a patient-criteria pair in our dataset and the color of the dot denotes its ground truth label (fail/success).

## Discussion and Conclusions

Clinical trials are a vital aspect of drug development but suffer from the time-intensive and expensive patient recruitment process. This work proposes a deep learning-based patient-trial matching framework, Siamese-PTM, that automatically predicts a patient’s eligibility for a clinical trial based on the criteria information. It involves training two identical encoders in tandem to effectively capture the complex semantics in the EHR and criteria data, while learning the correlations between them via the sharing of weights. The paired training samples in Siamese-PTM facilitate implicit data augmentation, allowing it to perform well even under a limited labeled data environment.

To reconcile the complication of matching complex criteria to the heterogeneous EHR, we inspect the informativeness of structured and unstructured EHR data on Siamese-PTM’s performance as independent and integrated feature sets through five input combinations. Unstructured EHR in the form the free-text radiology reports perform better than the structured EHR composed of clinical events. This finding emphasizes that unstructured narrative data contain more nuanced information that could provide discriminative cues for Siamese-PTM to accurately differentiate between eligible and non-eligible patients. The elaborate details of the patient’s health trajectory present in the unstructured EHR provide sufficient context for the LLaMA 2 pretrained model to learn semantically-rich EHR representations. Structured EHR, on the other hand, usually lacks this context as the clinical events represent data in a more concise format.

We develop a baseline rule-based classifier that relies on SQL-based feature engineering to perform the patient-trial matching task. Compared to the baseline results, Siamese-PTM achieved substantial performance improvement (40%). This highlights the feasibility of deep learning as a more convenient and effective alternative for data modeling, that can automatically learn the relevant features in the patient-trial data without manual labor and yield better performance. We evaluate Siamese-PTM in the coarse-grained and fine-grained settings, which result in the best F1 performance of 0.79 for the former and 0.92 for the latter. This indicates a 16% improvement achieved with token-level LLaMA 2 embeddings and promotes the importance of sequential modeling to enrich the feature learning of patient-trial data. Overall, Siamese-PTM consistently outperformed the Single-encoder models with performance gains ranging between 1%-20%. The early data fusion approach employed in Single-encoder models integrates features from the EHR and criteria data to a single input before the training phase that fails to efficiently capture their complementary semantics. Incorporating the data fusion in the training pipeline (intermediate data fusion) instead helps Siamese-PTM to successfully consolidate the correlating features from both EHR and criteria to learn a more meaningful representation. Comparisons of different loss functions to train Siamese-PTM revealed that the weighted variant of the standard binary cross-entropy loss (WBCE) helps to tackle the class imbalance in the patient-trial data, leading to better performance across most encoders.

This work has some limitations that we would like to address as future directions. The clinical trials used in this study mainly cover cognitive conditions, which limits Siamese-PTM’s generalizability to more diverse trials. Along similar lines, we evaluate Siamese-PTM on EHR data sourced from a single institution. Adapting Siamese-PTM for cross-institutional patient-trial matching is, however, a more challenging setting owing to the inherent domain shift in EHR data across different institutions.

## Data Availability

Protected Health Information (P HI) restrictions apply to the availability of the clinical data here, which were used under IRB approval for use only in the current study. As a result, this dataset is not publicly available. Qualified researchers affiliated with the Mayo Clinic may apply for access to these data through the Mayo
Clinic Institutional Review Board.

## Acknowledgements

This study is supported by the National Institute of Health (NIH) NIGMS (R00GM135488) and NIH R01AG084236.

